# Adhesion to VCAM1 and P-selectin Predict Time-to-Resolution (TTR) of Vaso-Occlusive Crisis

**DOI:** 10.1101/2022.10.20.22281335

**Authors:** Michael Tarasev, Xiufeng Gao, Jennell White, Marta Ferranti, Patrick C. Hines

## Abstract

Sickle cell disease (SCD) is characterized by frequent and unpredictable vaso-occlusive crises (VOCs) resulting in increased morbidity and mortality. Reliable biomarkers that predict the onset and progression of VOCs in SCD are unavailable, thus the existing standard of care is more focused on VOC intervention as opposed to VOC prevention. Sickle blood cells contribute to VOCs by adhering to the endothelium and aggregating to other blood cells in the circulation through pathologic adhesive interactions. In our previously described ELIPSIS study, blood samples were collected from 35 study subjects with SCD every 3 weeks during self-reported baseline and during self-reported VOCs (at home or in a healthcare setting). An electronic, patient-reported outcomes (ePRO) tool captured daily pain, VOC, and VOC resolution. Flow adhesion of whole blood to VCAM-1 (FA-WB-VCAM) and P-selectin (FA-WB-Psel) were assessed during each visit. Time-To-Resolution was established as the duration between the onset and subject self-reported resolution of VOC resolution and varied between 2 and 48 days. For the subset of TTR limited to ≤ 7 day, TTR was negatively correlated with FA-WB-PSel measured at the onset of VOC (R^2^=0.45; r=-0.67; *p*<0.05). Coefficient of determination increased to 0.62 when baseline FA-WB-VCAM levels were used as a second predictor in the multi-parametric model. In such a model, baseline FA-WB-VCAM was positively correlated with TTR at ≤ 7 day, with the difference in the sign of P-selectin and VCAM-1 effect on the reduction of pain (TTR duration) likely reflecting different mechanisms driving VOCs. Supplementation of FA-WB-VCAM and FA-WB-Psel multiparametric model with select blood chemistry biomarkers including several inflammatory mediators, further enhanced models ability to predict TTR. This study indicates that functional biomarkers obtained both at baseline and at the time of VOC can give insight into the time it may take for that specific VOC to resolve. These could assist providers in predicting which VOCs may require more intensive intervention. These data may also identify specific VOC phenotypes, allowing providers to intervene with a more patient-specific approach. Future studies are required to determine if FA-WB-VCAM and FA-WB-Psel can be used clinically to enable a more precision medicine-based approach to manage VOCs and if such an approach could result in improved outcomes and reduced healthcare costs by predicting VOCs for early intervention.

## Introduction

Sickle cell disease (SCD) is one of the most common genetic diseases. The pathologic hallmarks of the disease include red blood cell (RBC) sickling, chronic hemolysis, and increased erythrocyte adhesiveness to vascular endothelium that promote vaso-occlusion [1]. Vaso-occlusive events, including acute vaso-occlusive painful crisis (VOC), are major contributors to emergency department (ED) visits and hospitalizations. Based on the CDC 2015 survey, nearly half of all ED visits and about 75 percent of hospital admissions for SCD patients had a VOC diagnosis code [2]. Overall, 70 percent of SCD-related stays had a principal SCD diagnosis, and one-third of stays had a secondary diagnosis as SCD pain crisis [3].

Recurrent VOC has been associated with osteonecrosis leading to chronic arthritis, retinopathy, renal dysfunction, silent stroke, and end organ disease, with clinical progression resulting in shortened patient’s lifespan. VOC treatment typically involves nonspecific and supportive interventions such as intravenous hydration, RBC transfusions (simple or exchange), and analgesics (opioids, Toradol, acetaminophen, etc)that commensurate with the level of patient-reported pain. Patient-reported pain, a key metric in SCD diagnosis, positively correlates with high somatic symptom burden, health care utilization, effective coping, and opioid use [4]. However, due to its inherent subjectivity, measuring pain or relief of pain in VOC has been challenging to standardize both at home and during hospitalizations.

While VOCs often require urgent medical attention (e.g., ED visits and possible hospitalization), many, if not most, episodes are managed at home, without an ED visit, irrespective of severe pain [5, 6]. This issue is further complicated by the under-reported and under-appreciated prevalence of chronic pain accompanying the disease, with over half of SCD patients experiencing pain on more than half of the days and nearly a third of patients reporting pain on 95% of days [5]. Thus, there is a pressing need for objective criteria, based on clinically validated biomarkers, to predict, identify, and monitor acute painful crisis to provide guidance for VOC management and pain events in general.

Exact mechanisms involved in VOC have not been fully elucidated, however, inflammation, stress, increased viscosity, and hemolysis has been shown to promote endothelial activation and abnormal adhesive interactions to vascular endothelium [1]. Vascular occlusion starts through capillary clogging by cell-cell aggregates comprised of hemoglobin-S containing red blood cells (SS-RBC), leukocytes, and platelets. Subsequent increase in RBC transit time through the capillary network exacerbates ischemia, resulting in increased S type hemoglobin (HB-S) polymerization and SS-RBC sickling, thus causing a positive feed-back loop culminating in VOC [7]. Interactions between SS-RBC, platelets, leukocytes play a pivotal role in developing cell adhesion to activated endothelium. This adhesive cascade in initiated by P-Selectin mobilization on both activated platelets and endothelium cells and is essential in generating an inflammatory response [8, 9]. Additionally, P-selectin has been implicated as a mediator of normal and to a greater extent sickle RBCs to the endothelium [10]. There is also a body of evidence supporting the importance of additional adhesive interactions in VOC development and progression, including the of adhesive receptor very late antigen-4 (VLA-4) and its endothelial ligand vascular cell adhesion molecule 1 (VCAM-1) [11, 12].

We previously established that our proprietary, flow-based adhesion bioassays, whole blood (WB) adhesion to VCAM-1 (FF-WB-VCAM) and WB adhesion to P-selectin (FF-WB-Psel), stratifies SCD patients according to their risk of recurring VOCs or worsening of disease severity. We also demonstrated that baseline FF-WB-VCAM levels stratifies SCD patients based on disease severity (high or moderate/normal adhesion phenotype). In a 2-year prospective study, the high adhesion cohort experienced significantly more VOCs when compared to the normal/moderate adhesion phenotype [13]. In this study, we investigated additional relationships between FF-WB-VCAM and FF-WB-Psel, along with other chemical biomarkers, and the time to the resolution (TTR) from VOC in SCD subjects.

## Methods

### Study Participants and Sample Collection

Participant recruitment and blood sample collection were performed as previously described [14]. The protocol was approved by the Wayne State University Human Investigation Committee Institutional Review Board. Briefly, baseline blood samples (non-VOC) were collected from SCD subjects (n=35) every three weeks in the participant’s home by mobile phlebotomy over a six-month time period. An electronic, patient-reported outcomes (ePRO) tool captured daily pain, VOC, and VOC resolution. Patient reported vaso-occlusive pain crisis (VOC) on the ePRO device triggered a series of blood collections: within 24- and 48-hours (blood sample 1 and 2, respectively) and following VOC resolution (sample 3; patient reporting non-VOC state for two consecutive days).

### Clinical Biomarkers

Whole blood samples were sent to Myriad RBM (Austin, TX) or Detroit Medical Center to measure complete blood counts, hemoglobin differentiation and a multiplex panel of analytes that included: percentage and absolute number of reticulocytes; Uric Acid; Interleukins (IL) 1A, 1B 1RA, 2, 3, 4, 5, 6, 7, 8, 10, 15, 17, 18, and 23; soluble suppression of tumorigenicity 2; subunits of the Interleukin 12 family cytokines (IL12P40 and IL12P70); Interleukin-6 receptor subunit alpha (IL6R); interleukin-33 receptor (IL33R); Lactate Dehydrogenase (LDH); Brain-Derived Neurotrophic Factor (BDNF); high affinity scavenger receptor for the hemoglobin-haptoglobin complex Cluster of Differentiation 163; Granulocyte-macrophage colony-stimulating factor (GM-CSF); Growth/Differentiation Factor-15 (GDF-15); Stem Cell Factor (SCF); Tissue Inhibitor of Metalloproteinase 1 (TIMP1); Tumor Necrosis Factor α (TNFα); Tumor necrosis Factor β (TNFβ); Tumor necrosis factor receptor 2 (TNFR2); Vascular Endothelial Growth Factor (VEGF); Factor VII (FVII); Factor XII (F12); Thrombocyte Agglutination test; Tissue Factor III (TF3); Alanine Transaminase (ALT); Aspartate Aminotransferase (AST); Intercellular Adhesion Molecule 1 (ICAM-1); Interferon gamma (IFN-γ); Interferon-Inducible Protein 10 (IP10); Macrophage inflammatory protein-1α (MIP1A); Macrophage inflammatory protein-1β (MIP-1B); Macrophage Inflammatory Protein-3α (MIP3A); the Matrix Metalloproteinase 3 (MMP3) and Matrix Metalloproteinase 9 (MMP9); Monocyte chemotactic protein (MCP) 1, 2 and 4; Myeloid progenitor inhibitory factor 1 (MPIF1); a selective attractant protein for memory T lymphocytes and monocytes, “Regulated on Activation, Normal T Expressed and Secreted” (RANTES); total plasma protein concentration (albumin, gamma-globulins, etc.) (PPC); serum concentrations of albumin, myoglobin, bilirubin, ferritin, creatinine, Ca^++^, CO_2_, Cl^-^, and glucose; Osteocalcin, also known as bone gamma-carboxyglutamic acid-containing protein; Periostin, a matricellular protein that mediates cell activation by binding to receptors present on the cell surface; the difference between the negatively charged and positively charged electrolytes in the blood; D-DIMER; C-reactive protein (CRP); Adiponectin, a protein produced and secreted by fat cells; alpha-2-macroglobulin (α2MG); the extracellular newly identified receptor for advanced glycation end-products binding protein (EN-RAGE); Eotaxin 1, an important eosinophil-specific chemokine; Soluble Vascular Cell Adhesion Molecule 1 (VCAM-1); Soluble E-selectin; Soluble P-selectin; Plasminogen activator inhibitor type 1 (PAI1); blood urea nitrogen (BUN) test; Monokine Induced by Gamma Interferon (MIG); the estimated glomerular filtration rate (eGFR). These biomarkers were previously proposed as candidate biomarkers that were either elevated in SCD subjects during stable, non-VOC days [15, 16], or elevated in patients experiencing a VOC as compared with a separate cohort of non-VOC control subjects [17]. To distinguish the values obtained at baseline from those obtained at VOC, biomarkers were marked a “-B” or a “-V” index referring to Baseline or VOC, respectively.

### Flow Adhesion Biomarkers

Standardized microfluidic flow adhesion assays, including flow adhesion of whole blood to VCAM-1 (FA-WB-VCAM) and flow adhesion of whole blood to P-selectin (FA-WB-Psel) were conducted as described previously [12]. Briefly, the assays were performed using a commercial well-plate microfluidic flow adhesion system, BioFlux 1000Z (Fluxion Bio, San Francisco, CA). Whole blood samples (1:1 diluted with HBSS buffer) were perfused through VCAM-1 or P-selectin coated microfluidic channels using pulsatile (1.67Hz) flow at 1 dynes/cm^2^, and then washed at the same conditions to eliminate non-adherent cells. Images were acquired with a high-resolution CCD camera and analyzed with BioFlux Montage imaging software (Molecular Devices, Downingtown, PA). An adhesion index (AI) was established for each sample by quantifying adherent cells within a standard viewing area (cells/mm^2^).

### Data Processing and Analysis

Biomarker data were categorized into VOC (marked with -V), and baseline (marked with -B) and separated into four timeframes based on the time between the onset of patient-reported VOC and the Time to Resolution (TTR) of the corresponding VOC. Selected timeframes were ≤ 7 days (TTR_1_), ≤ 14 days (TTR_2_), ≤ 21 days (TTR_3_), and up to 49 days (TTR_4_). Simple and multi-parametric linear mixed regression models accounted for repeated measurements in independent subjects and were used to assess the predictive value of biomarkers described above. At ≤ 7 days (TTR_1_), only one subject reported the resolution twice, in which case, the values were averaged between the two events for simplified analysis. Models were also compared by Kaplan-Meier estimates and log-rank test to allow risk stratification based on “survival” (times-to-event) between patient’s groups established through biomarkers. Data were analyzed using R statistical software, version 4.0. (R Foundation for Statistical Computing, Vienna, Austria); *p*-values <0.05 were considered statistically significant.

## Results

The distribution of TTR durations (TTR_1_, TTR_2_, TTR_3_, and TTR_4_) for the VOC events recorded in the study is shown in Figure 1. TTR_1_ (durations at ≤7 days) significantly correlated with FA-WB-Psel measured during VOC (Psel-V; Figure 2). No correlation was observed between TTR_1-4_ and FA-WB-VCAM during VOC (VCAM-V), or FA-WB-Psel and FA-WB-VCAM measured at baseline (Psel-B and VCAM-B, respectively). Select chemical biomarkers at baseline and during VOC significantly correlated with TTR_1_ (Table 2 and Figure 3). Additionally, Hb and MIG at baseline and EN-RAGE, MMP3, IL6R, Absolute Monocyte Count (AMC) and monocyte percentage (MONOCYTE) during VOC approached significance at 0.05<p<0.1 and were considered in multi-parametric model analysis. No significant correlations were observed between biomarkers (described above) and TTR longer than 7 days (TTR_2-4_). Age and gender, and all other biomarkers did not correlate with TTR at any timeframe (TTR_1-4_).

**Table 1.**
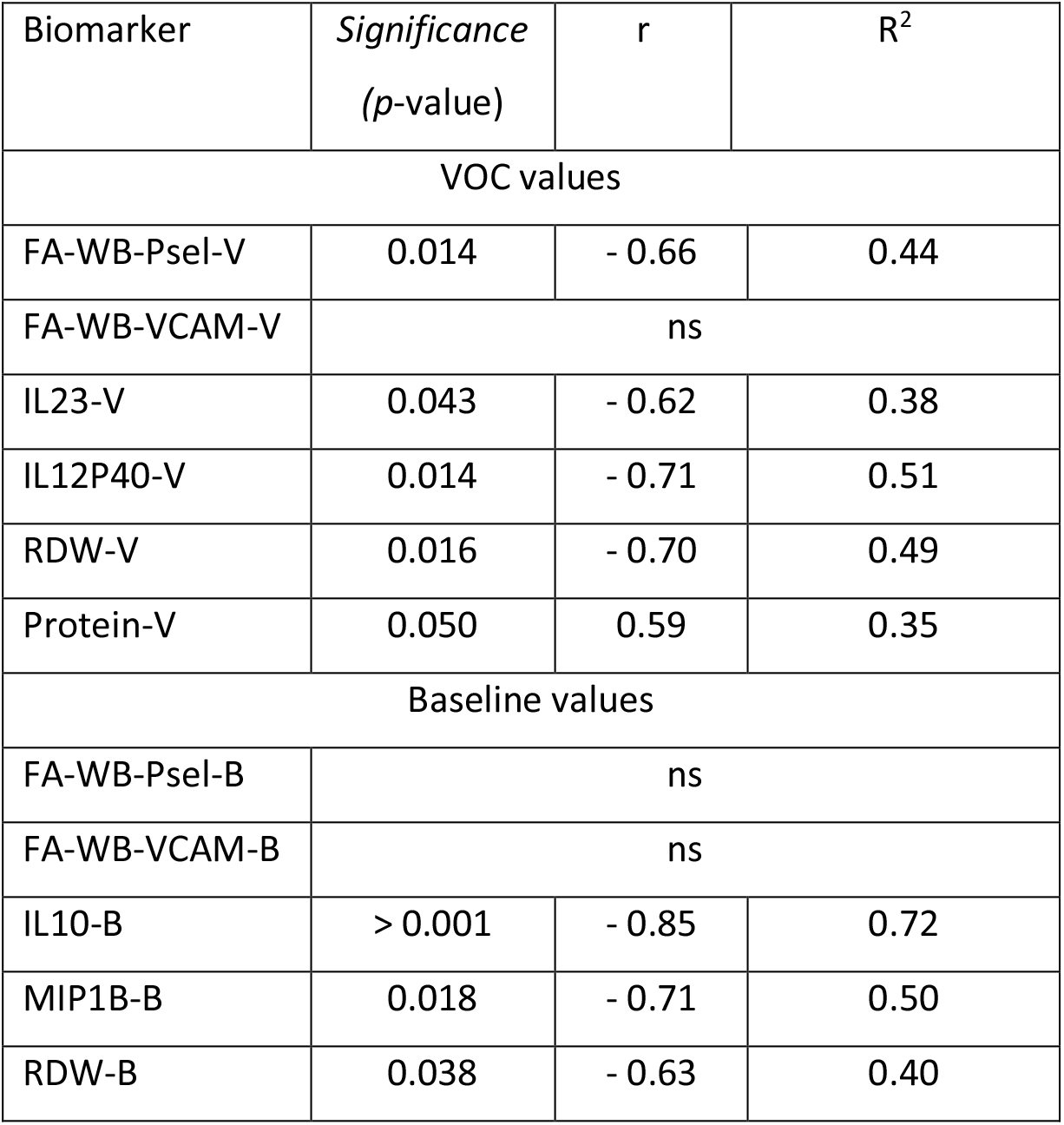
Single biomarker correlations to Time-to-Resolution (TTR ≤ 7 days)

**Table 2.** Coefficients of determination in a combination two independent variables model predicting TTR_1_ (Time-to-Resolution ≤ 7 days) including clinical chemistry markers with statistical significance

| A<sup>1</sup> | Statistically significant |  |  |  |  |  |  |  | ns |
| --- | --- | --- | --- | --- | --- | --- | --- | --- | --- |
|  | IL12P40-V | IL23-V | RDW-V | PROT-V | IL10-B | MIP1B-B | RDW-B | PSel-V | VCAM-B |
| IL12P40-V | **0.51** | ns | **0.61** | **0.62** | ns | ns | **0.65** | ns | ns |
| IL23-V |  | **0.38** | ns | ns | ns | ns | ns | ns | ns |
| RDW-V |  |  | **0.49** | **0.72** | ns | ns | ns | ns | ns |
| PROT-V |  |  |  | **0.35** | **0.83** | **0.52** | **0.54** | ns | ns |
| IL10-B |  |  |  |  | **0.68** | ns | **0.71** | ns | ns |
| MIP1B-B |  |  |  |  |  | **0.42** | **0.64** | **0.58** | ns |
| RDW-B |  |  |  |  |  |  | **0.45** | ns | ns |
| PSel-V |  |  |  |  |  |  |  | **0.45** | **0.62** |
| VCAM-B<sup>2</sup> |  |  |  |  |  |  |  |  | **0.03** |

<sup>1</sup>Key: **0.51** - all biomarkers in the model are statistically significant at p<0.05 and the model is significant at F <0.05; **0.69** - one biomarker is significant at p<0.05 and one biomarker is approaching significance at 0.5<p<0.1, and the model is significant at F <0.05; ns – no significance.
<sup>2</sup>VCAM-B (WB-FA-VCAM at baseline) by itself shows low predictive capability towards TTR<sub>1</sub>

**Figure 1.**
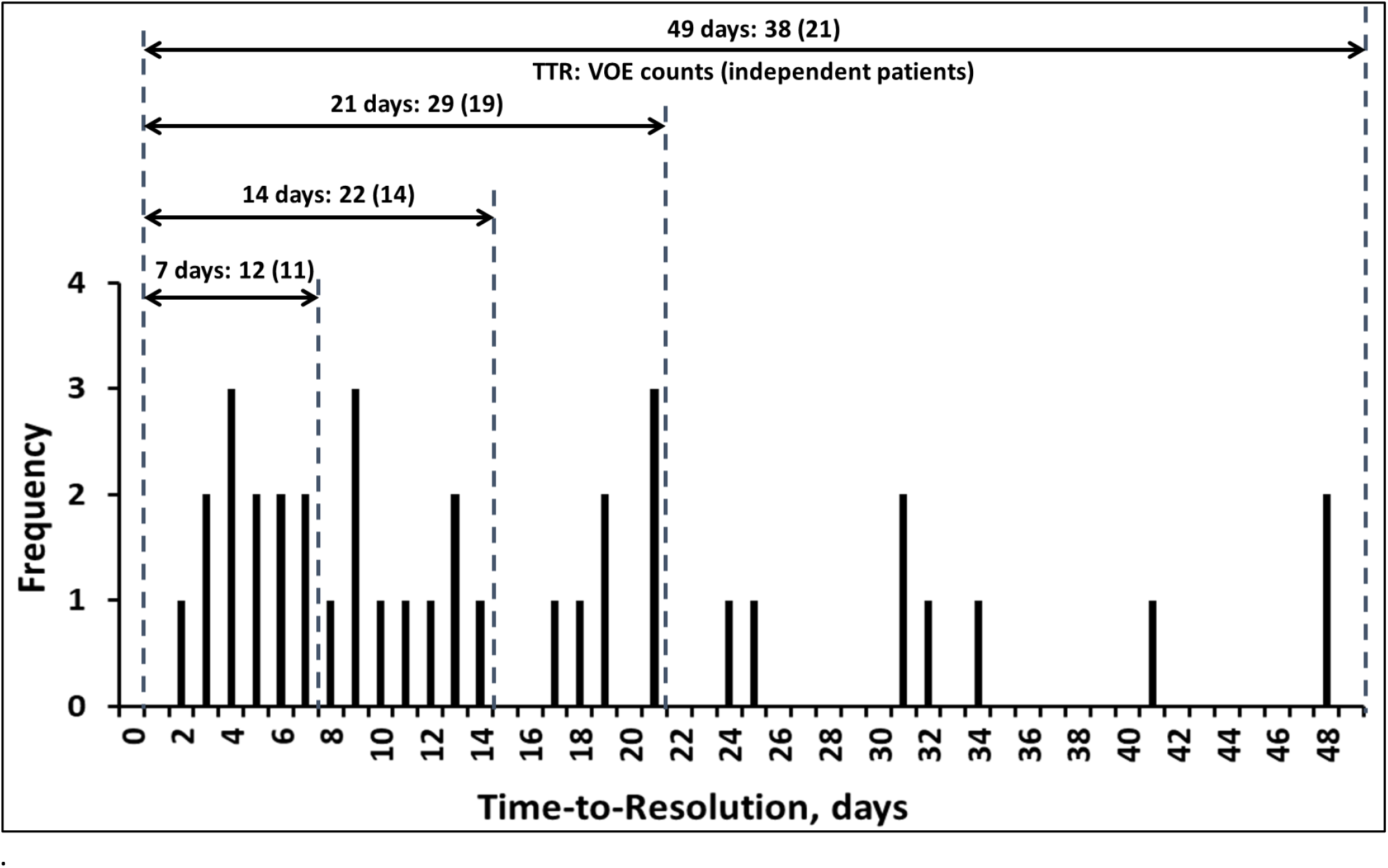
Time-to-Resolution duration for the 38 VOC events recorder in the study. 7-day, 14-day, and 21-day, and 49-day intervals are shown with corresponding count of independent patients and associated VOC events.

**Figure 2.**
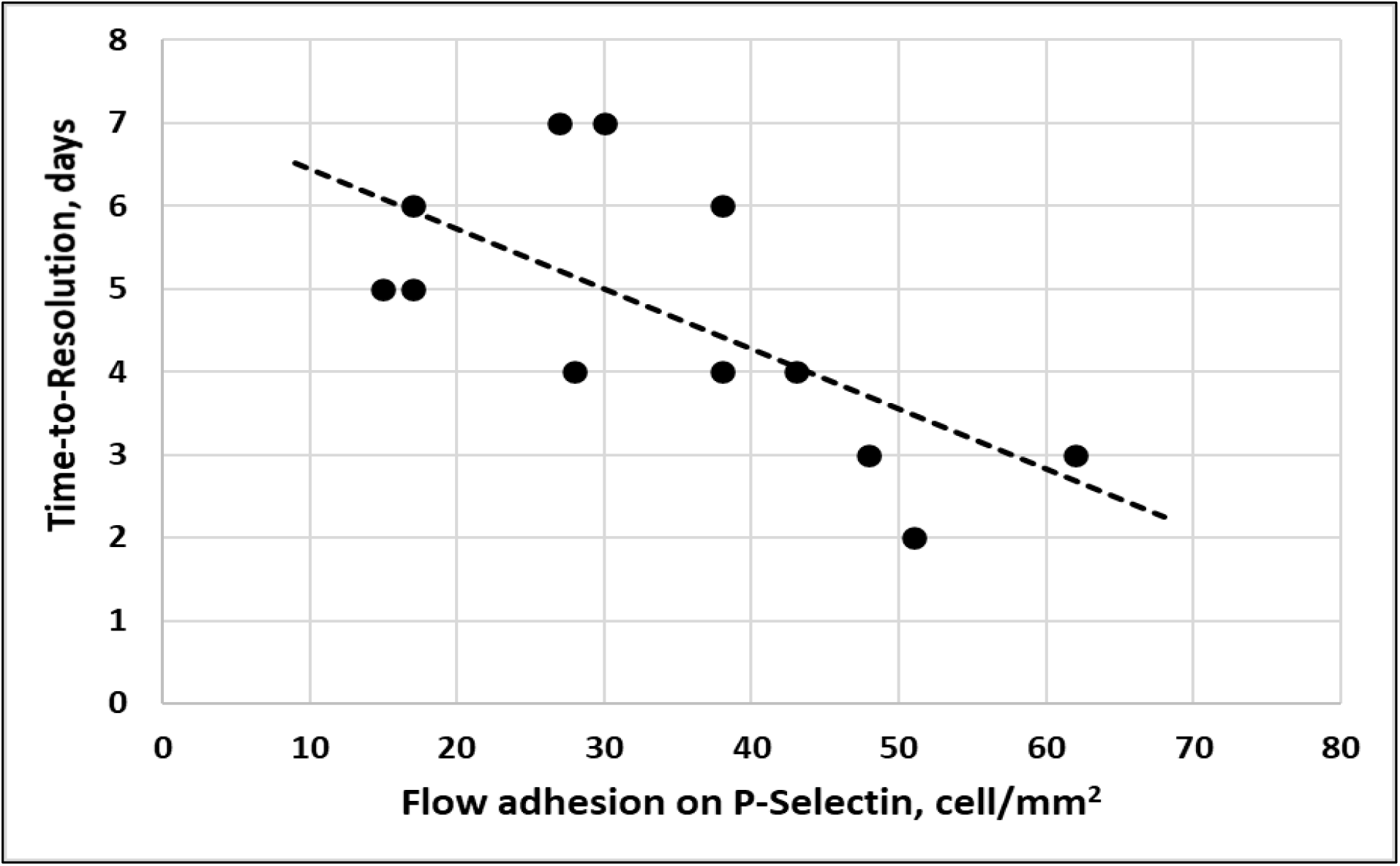
Time-to-Resolution at seven days or less as a function of WB-FA-Psel measured at VOC.

**Figure 3.**
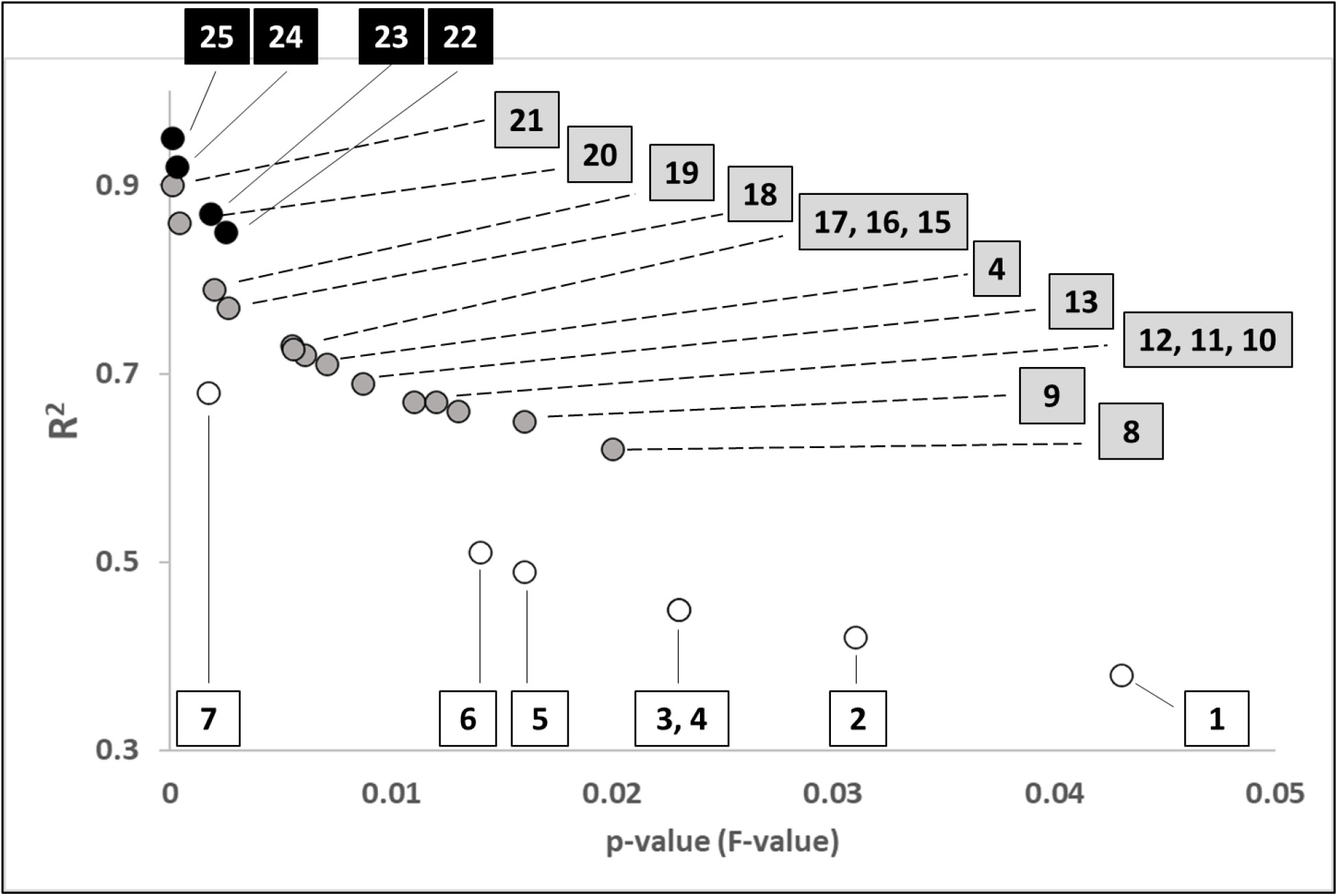
Predicting variables for TTR less or equal to 7 days using a single predictor variable (⚪): 1. IL23-V; 2. MIP1B-B; 3. RDW-B; 4. PSel-V; 5. RDW-V; 6. IL12P40-V; 7. IL10-B;); two predictor variables (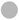): 8. PROTEIN-V and MIG-B; 9. IL23-V and BUN-B; 10. MIP1B-B and PSel-V; 11. MIP1B-B and IL6R-V; 12. MIG-B and IL23-V; 13. PSel-V and VCAM-B; 14. MIP1B-B and RDW-B; 15. IL12P40-V and RDW-B; 16. En-Range-V and RDW-V; 17. En-Rage-V and RDW-V; 18. PROTEIN-V and RDW-V; 19. IL6R and IL12P40-V; 20. IL10-V and PROTEIN-V 21. IL10-B and En-Rage-V; and three predictor variables (●): 22. IL23-V; En-Rage-V, and MIG-D; 23. PSel-V, VCAM-B, and MIP1B-B; 24. IL10-B, RDW-V, and PROTEIN-V; 25. IL10-B, En-Rage-V, and VCAM-B. Significance is shown as *p-value* for a single predictor and *F-value* for two or three predictors.

Models for VOC risk-stratification assessed at TTR_1-4_ were compared to Psel-V and VCAM-B using Akaike information criterion (AIC) with lower AIC values corresponding to higher model quality. For both biomarkers, VOC risk-stratification weakened with increased TTR duration (TTR_1-4_; Figure 4). TTR_1_ represented the lowest AIC values averaged across the range of cut-off AIC values: from 37 to 53 cells/mm2 for FA-WB-Psel-V (Psel-V) and from 360 to 542 for FA-WB-VCAM-B (VCAM-B). The ranges were based on the previously established baseline adhesion cut-off values for VOC risk stratification (46 cells/mm^2^ for Psel-V and 408 cells/mm^2^ for VCAM-B) [18]. For both biomarkers changes in AIC at TTR_1_ were too small to allow for meaningful optimization of the cut-off interval. Hazard Ratio (HR) values indicated a potentially higher impact of Psel-V on TTR, than of VCAM-B (HR of 13 and 0.36 correspondingly) and is consistent with higher VCAM-B and lower Psel-V values being associated with longer TTR. The 95% confidence intervals for HR values calculated for TTR_1_ were very broad (2.1-79 and 0.093–1.4 correspondingly) which combined with the small sample size does not allow for meaningful risk stratification based on HR values.

**Figure 4.**
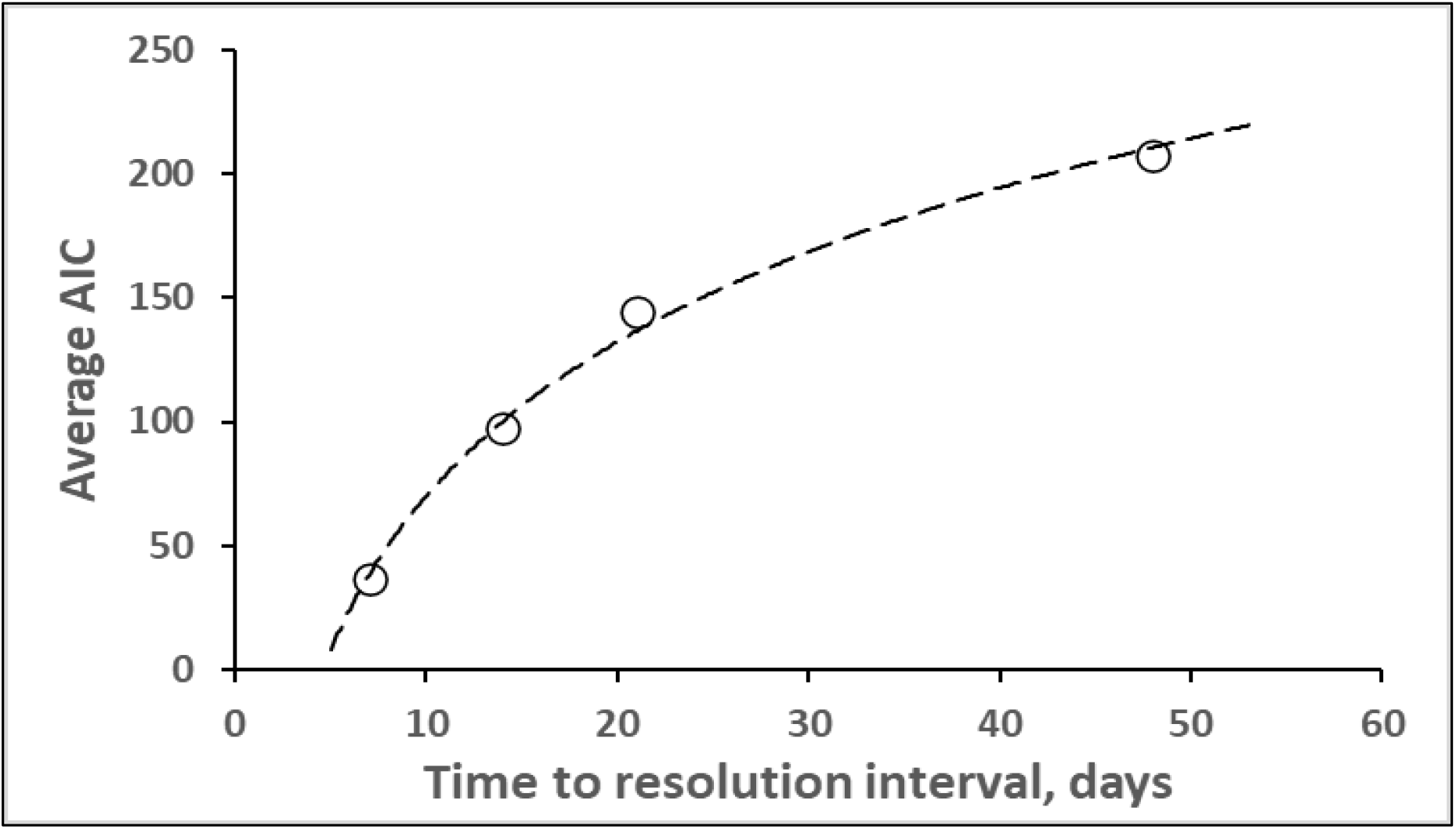
Average AIC values as a function of the selected TTR interval. Logarithmic function fit is for illustrative purpose only.

Multi-variate analyses assessed biomarker combinations, as compared to single-value biomarkers, in their ability to potentially increase predictive capability of the model toward TTR_1_ (≤7 days). In addition to flow adhesion biomarkers, clinical chemistry markers showing independent significance in prediction of variance in TTR duration were also evaluated for inclusion in multi-parametric models. Cross-correlative analysis demonstrated that FA-WB-Psel-V positively correlated with RBC distribution width (RDW) (p<0.01, R^2^=0.57), IL10 (p<0.01, R^2^ =0.59) at baseline, and AMC (p<0.01, R^2^=0.65) and IL6R-V (p=0.04, R^2^=0.4) during VOC, and negatively correlated with Hb (p = 0.04, R^2^=0.4) at baseline (Figure 3).

Coefficients of determination, representing proportion of TTR_1_ variance explained by a combination of two biomarkers, each independently statistically significant, are presented in Table 2. Considering the limited sample size of TTR_1_, also evaluated were clinical chemistry markers with the potential for significance (p-values between 0.05 and 0.1). Coefficients of determination, representing proportion of TTR variance explained by a linear combination of two biomarkers, including one with potential for significance, are given in Table 3. Despite lacking independent significance, inclusion of VCAM-B as a secondary biomarker when combined with many independently significant clinical chemistry markers, significantly improved the predictive capability of the two-biomarker model. When in combination with Psel-B, inclusion of VCAM-B significantly improved the two-biomarker model with both adhesion biomarkers (Psel-V and VCAM-B) reaching significance and with model F-significance < 0.01, and R^2^ values increased to 0.69 from R^2^ =0.44 observed for FA-WB-Psel-V alone (Table 2 and Figure 3). Signs of model coefficients indicate that shorter TTR, in the 2-to-7-day time interval, are associated with increased adhesion to P-selectin during VOC (Psel-V) and decreased RBC adhesion to VCAM-1 at baseline (VCAM-B). Of the other biomarkers showing statistical significance in a 2-marker combination (Tables 2 and 3), all were negatively correlated with TTR, except for PPC and EN-RAGE biomarkers during VOC that positively correlated with TTR in all biomarker combinations.

**Table 3.** Coefficients of determination in a combination two independent variables model predicting TTR_1_ (Time-to-Resolution ≤ 7 days) including clinical chemistry markers approaching statistical significance

| **B<sup>1</sup>** | **Approaching statistical significance** |  |  |  |  |  |  |
| --- | --- | --- | --- | --- | --- | --- | --- |
|  | **Hb-B** | **ENRAGE-V** | **BUN-B** | **MIG-B** | **MMP3-V** | **IL6R-V** | **AMC-V** |
| **IL12P40-V** | **0.60** | ns |  | **0.58** | ns | **0.74** | ns |
| **IL23-V** | ns | ns | **0.56** | **0.58** | ns | **0.47** | ns |
| **RDW-V** | ns | **0.66** | ns | ns | **0.48** | ns | ns |
| **PROT-V** | ns | ns | ns | **0.53** | ns | ns | ns |
| **IL10-B** | ns | **0.88** | ns | ns | ns | ns | ns |
| **MIP1B-B** | ns | ns | ns | ns | ns | **0.59** | **0.52** |
| **RDW-B** | ns | ns | ns | **0.60** | ns | ns | ns |
| **PSel-V** | ns | **0.55** | ns | **0.57** | ns | ns | ns |
| **VCAM-B** | ns | ns | ns | ns | ns | **0.43** | ns |

<sup>1</sup>The key is the same as in Table 2.

Survival analysis did not show a statistically significant model when using Psel-V and VCAM-B as a 2-parameter set of independent predictors of TTR_2-4_. Like in the single parameter models, AIC values were lowest at up to 7-day TTR duration, as compared to 14, 21 or 48 days, indicating that use of 7-day TTR (TTR_1_) allows for the most reliable model. For a given TTR_2-4_ duration, AIC changes were too small to allow for reliable optimization of Psel-V and VCAM-B cut-off values.

Significance of the results with a 2-biomarker model was markedly improved when the analysis was performed with the integrated adhesion biomarker IFA metric calculated based on regression analysis using Psel-V and VCAM-B as the combined predictors of TTR duration (Formula 1). Correlation of TTR_1_ with the IFA biomarker is presented on Figure 5. (Correlation coefficients of the linear regression model were −0.0734 for Psel-V and 0.0044 for VCAM-B). With the intercept of 5.78, IFA metric represents TTR duration as predicted by the model (Formula 1) based on the data from the events resolved within one week (TTR_1_).

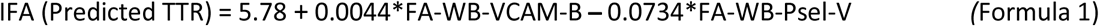

**Figure 5.**
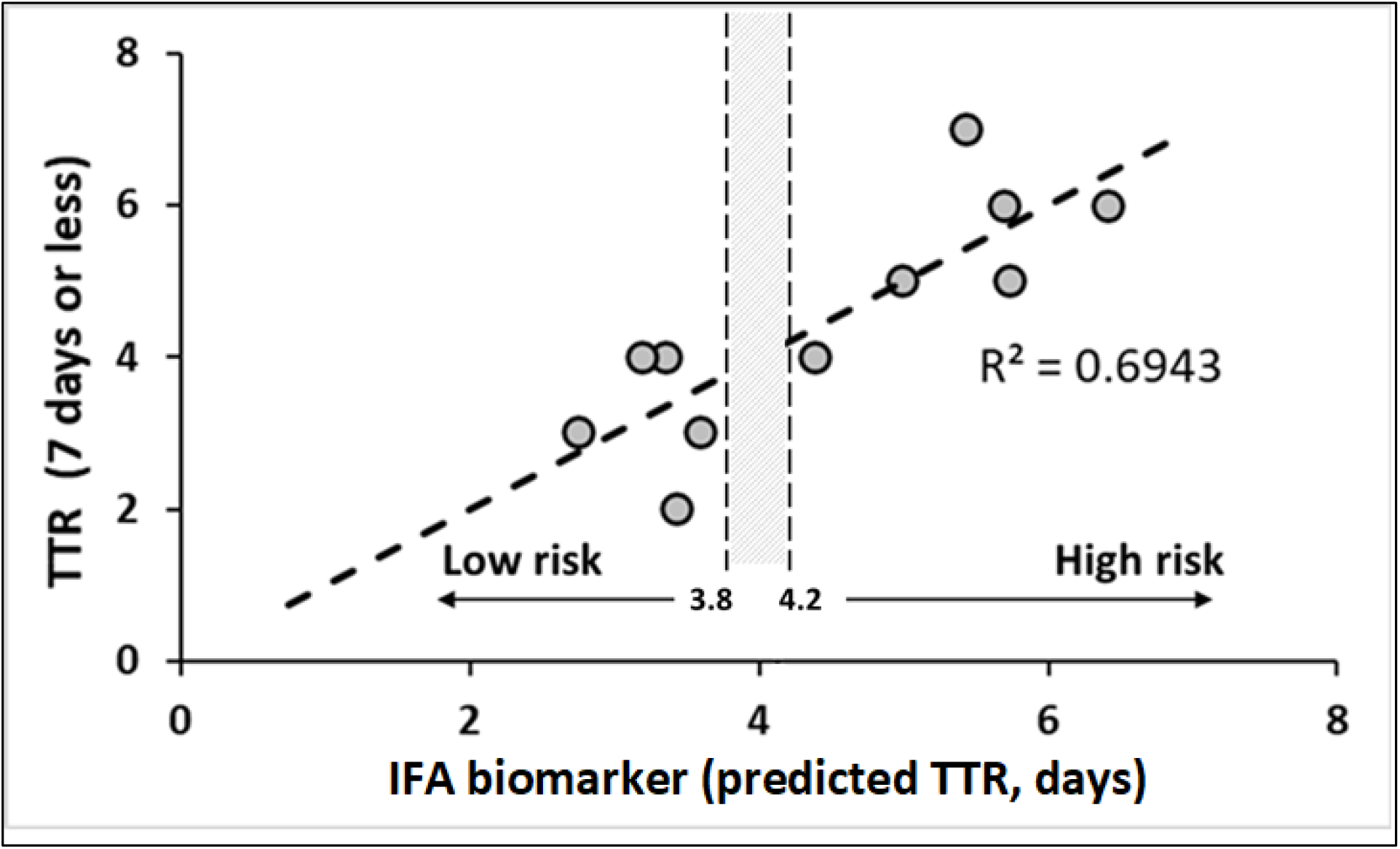
Time to Resolution at seven days or less (TTR_1_) as a function of an integrated flow adhesion biomarker (IFA) based on WB-FA-Psel measured at VOC (PSel-V) and WB-FA-VCAM measured at baseline (VCAM-B). IFA values were calculated using Formula 1.

Optimization of the model for TTR durations not exceeding 7 days through variation of cut-off values showed significant (over 8 AIC units) reduction in AIC values, with AIC minimum located in the range of 3.7 – 4.2 for IFA values (Figure 6). Within the optimal IFA range for that model, HR value was 0.055 (95% CI 0.006 – 0.51) indicating potential utility of IFA metric for stratification of VOC patients based on anticipated time to recovery from the crisis. Kaplan-Meier plot using the optimized IFA value is presented on Figure 7. The model was based on 12 events with average TTR of 3.2 ± 0.3 days for 5 events and 5.7 ± 0.4 days for 7 evens (p < 0.01). More precise optimization, potentially leading to narrower HR confidence intervals, was not possible due to sample size limitations. Models based on longer TTR interval durations lacked significance.

**Figure 6.**
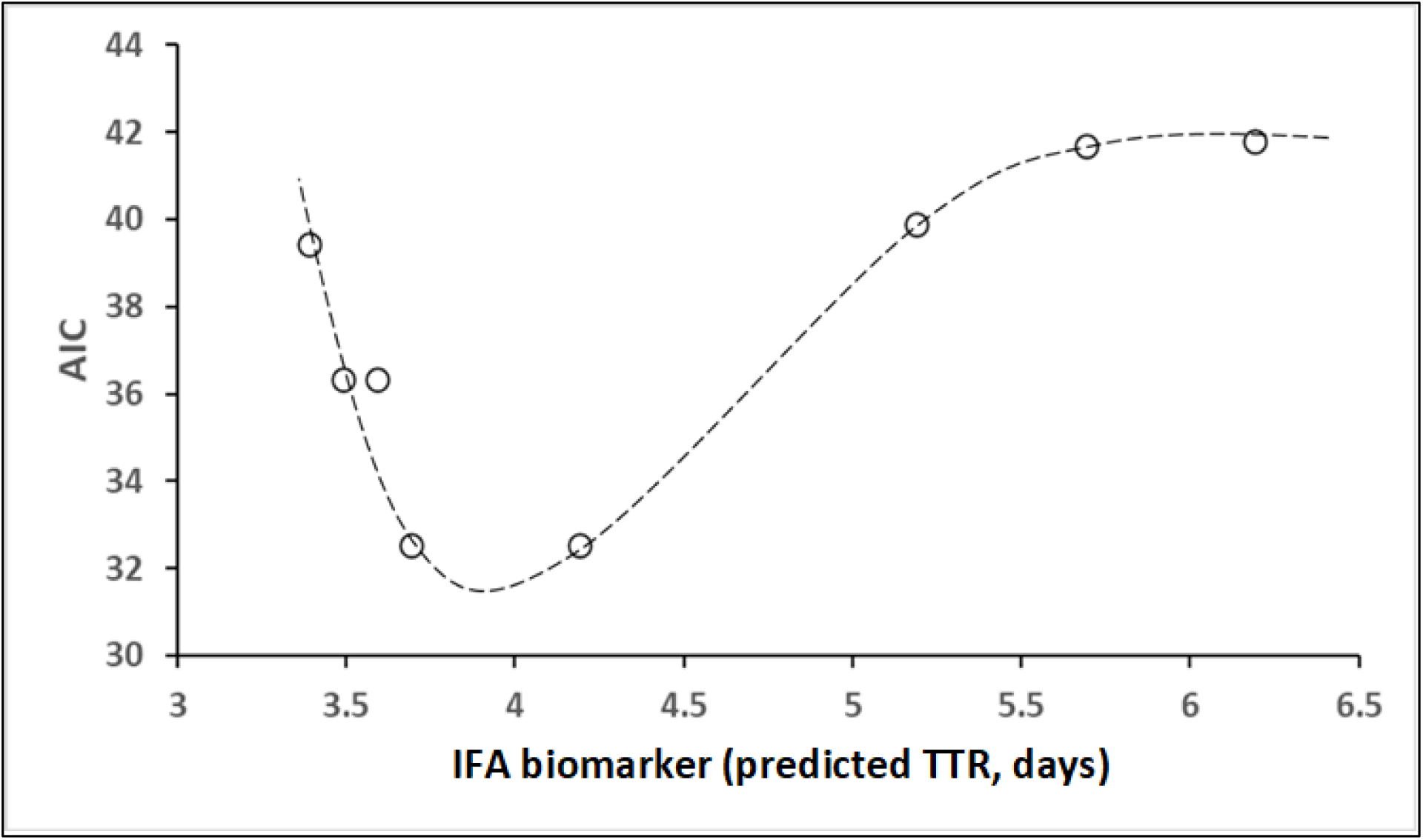
Changes in AIC as a function of the cut-off value of the integrated flow adhesion (IFA) biomarker. Fit is for illustrative purpose only. IFA values were calculated using Formula 1.

**Figure 7.**
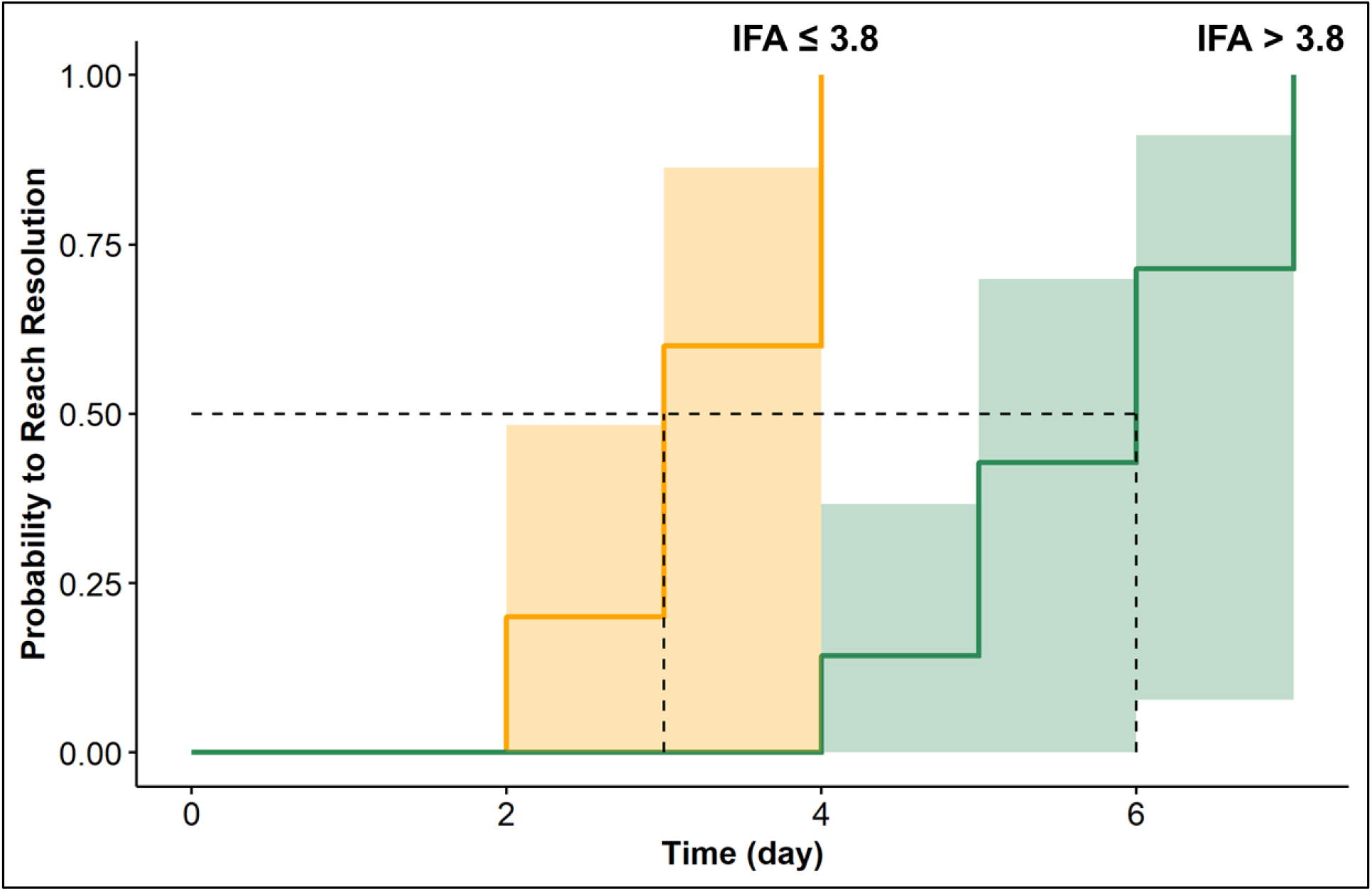
Kaplan-Maier plots for risk of extended TTR based on the integrated flow adhesion (IFA) metric calculated as a liner combination of flow adhesion on VCAM at baseline (VCAM-B) and on P-Selectin during the VOC (PSel-V) with the optimal IFA cut-off value of 3.8 determined through AIC minimization (Fig. 6).

Further increase in multiparametric model predictive capability can be achieved by supplementing the 2-marker models with additional markers showing independent significance in predicting TTR. Only four models, with all 3 parameters showing independent significance at p<0.05 had been identified (Figure 3). Of those two combinations include flow adhesion biomarkers with the highest model predictive capability (R^2^ of about 0.95) shown by a combination of VCAM-B and IL10-B values with EN-RAGE-V. High predictive capability (R^2^ of about 0.87) was also shown by a model combining FA-WB-Psel-V with VCAM-B and MIP1B-B values. Several other 3-parametric model with predictors that included IL23-V, EN-RAGE-V, MIG-B, IL10-B, RDW-V and PPC-V, showed significance (F-value<0.01, R^2^ > 0.75) despite intercorrelation between the predictors. These models have at least two predictors showing independent significance (p<0.05) with the third predictor lacking independent significance (p>0.05) but allowing for a marked (> 0.1 units) increase in correlation coefficient value (data not shown).

Over the 6-month study duration, 12 subjects experienced VOC more than once, which allowed the calculation of the “Time to the Next VOC Event” (TNE), defined as the duration between the self-reported resolution of one crisis and the onset of the next one (TNE: 54 ± 38 (mean ± SD) days, range 13 – 136 days). One subject experienced 4 VOC events (3 TNE values) and two subjects experiences 3 VOC events each (2 TNE values each). Analysis using repeated measurements model (implemented in R) did not reveal any statistically significant correlations between evaluated biomarkers and TNE durations. For datasets including shorter TTR durations (TTR_1_, TTR_2_), regression models using TTR as a predictor on TNE showed higher coefficients of correlations and lower p-values, however significance at p < 0.05 was not reached. Failure to establish significance could be due to rapidly declining sample size at shorter TTRs, which preclude rigorous evaluation.

## Discussion

Blood cell adhesion to the vascular endothelium is a crucial event in the pathophysiology of VOCs and other clinical manifestations of SCD [19]. VCAM-1/VLA-4 and P-Selectin/PSGL-1 interactions were reported as both independently and cumulatively contributing to risks of vaso-occlusive events [18, 20]. High levels of VLA-4 and VCAM-1 have been associated with vaso-occlusive adhesive events and severe disease phenotypes, and with a response to SCD-modifying therapy [21-26]. Elevated VLA-4/ VCAM-1 binding at baseline strongly correlates with SCD severity, providing an on-going, and relatively slow changing background for RBC-endothelial adhesive interactions [13].

Faster VOC resolution was strongly correlated with higher FA-WB-Psel during VOC (Psel-V), with no correlation observed with adhesion at baseline (Psel-B). These observations may demonstrate the role of fast transient elevation of P-selectin adhesion in the development and resolution of VOCs. Both VCAM-1/VLA-4 and P-selectin/PSGL-1 interactions contribute to cell adhesion to endothelium which promotes vaso-occlusion and ultimately VOCs. Associated VOCs induced by both VCAM-1 and P-selectin mediated cell adhesion to endothelium would be dependent on both slowly changing VCAM-1 baseline adhesion that may be associated with overall disease severity and acute and transient elevation of P-selectin adhesive interactions. Patients with lower background VCAM adhesion may require more significant contribution from P-selectin adhesion for significant vaso-occlusion, while a smaller increase in P-selectin adhesion may be required to initiate such events on a high VCAM adhesion background. High baseline P-selectin adhesion in this model would still promote elevated SCD severity, even without, based on this study results, association with VOC time to resolution. P-selectin, which upon activation is rapidly transferred from secretory granules to plasma membranes, can also be quickly internalized and degraded inside the cell [27-29]. Thus, VOCs that are induced by transient spikes in P-selectin adhesion may have a faster decline or reversal of a more transient P-selectin-mediated adhesive interactions to their pre-VOC levels, thus promoting faster recovery from VOC (shorter TTR). That is consistent with the negative correlation between P-selectin adhesion at VOC and TTR_1_ reported in this study.

In this work we also observed that faster TTR of VOCs correlated with lower values of FA-WB-VCAM during clinical baseline (VCAM-B), albeit the significance was reached only when in combination with PSel-V marker. No correlation was observed with FA-WB-VCAM measured during VOC. Previous reports suggested that FA-WB-VCAM at clinical baseline as one of the determinants of the overall disease severity [13], with such elevated severity potentially contributing to P-selectin-driven onset of VOC. Additionally, increased VCAM-1/VLA-4 interactions may lengthen TTR because more adhesive sickle RBCs may become trapped in post capillary venules, further contributing to VOC events. Microvascular occlusion through accumulation of sickle RBCs and other adherent cells in the absence of an inflammatory trigger has been reported previously [7]. VOC pain in this context may not resolve with anti-inflammatory therapy (e.g., NSAIDS), thus resulting in a subjectively longer TTRs.

In this study, TTR was significantly predicted by a combination of adhesion and clinical chemistry biomarkers when the VOCs resolved within 7-days. No significant associations were found for longer TTR durations (from 14 to 48 days), possibly because such TTR could be more indicative of the different resolution mechanisms driven by “chronic” or “neuropathic” pain. Uncertainty in the actual TTR duration combined with potential misdiagnosis of transiently exacerbated chronic pain upon VOC onset, represent one of the limitations of this study.

Hospital and ED VOC treatment were difficult to account for in this study design. A larger sample size would be desirable to better assess and validate the dependence of clinical adhesion and biochemical markers on TTR after VOC. Considering the limited sample size, correlations reported in this study should be considered as provisional, but indicative of biomarkers to target in the follow-up investigations. Selection of patients with significantly different probabilities of VOC events could help determine TTR dependence on disease severity. Additionally, all study subjects were recruited in southeast Michigan, and this may impact generalizability of the results.

It is known that SCD, particularly VOCs, are associated with elevated levels of proinflammatory cytokines and acute-phase proteins, as well as reduced levels of anti-inflammatory cytokines at baseline (in steady-state) and during VOC [30]. Specifically, levels of IL-6, a cytokine responsible for systemic features of acute pain response, were reported to be increased, and levels of IL-10, an anti-inflammatory cytokine downregulating IL-6, significantly reduced during VOC compared to steady-state. Increased IL-10 and decreased Il-6 levels at VOC were further reported to be correlated with shorter TTR durations [31]. In this study, baseline IL10 and MIP1, a macrophage protein serving as a chemoattractant for immune cells, negatively correlated with TTR, indicating that higher levels of these cytokines are likely associated with a more rapidly reversible acute inflammatory process, as compared to other VOC resolution mechanisms that may be less responsive to recovery time, anti-inflammatory meds, or other interventions.

During VOC, several cytokines (Tables 2 and 3) associated with pathogenic inflammatory processes and host immune response, were negatively correlated with TTR_1_ and may have contributed to promoting vaso-occlusion with a stronger immune response potentially leading to faster resolution of VOC. Monocyte and absolute monocyte counts negatively correlated with TTR_1_ indicating that higher counts result in a stronger immune response and a shortened recovery from vaso-occlusion.

In parallel, EN-RAGE, a proinflammatory protein secreted by activated granulocytes, macrophages, and lymphocyte, positively correlated with TTR_1_. This biomarker may not be the driver of VOC, but a result of an ongoing immune-derived inflammatory response. Elevated gamma-globulins that can be linked to inflammation and drive elevation of the PROTEIN biomarker, a potential result of past inflammatory action, a likely cause of VOC.

Characteristic nociceptive pain (primarily related to visceral and somatic-tissue injury) arises as a result of the sequelae of pathophysiologic SCD events during VOC that comprises inflammation and vaso-occlusion [32]. Consequently, subjectively experienced VOC pain may be supplemented pain due to persistent low-grade inflammation involving the chest, back, or upper and lower extremities associated with chronic pain syndromes [33]. Neuropathic pain remains poorly defined in SCD, but onic pain can complicate the determination of the time-to-resolution (TTR) both in hospital- and in self-reported scenarios.

Previously reported average VOC duration was 2.0 +/- 1.8 days [34] or alternatively 6-7 days were given as time to pain resolution [35]. The mean length of VOC-associated hospital stays was reported as 5-6 days for adults and 4-5 days for children [3, 36]. These discrepancies are due, in part, to uncertainties in defining “time-to-resolution” and its association with declining pain levels. Discrepancies could also arise, especially for longer TTRs, due to “resolution” being more reflective not of subsiding pain crisis, but of alleviation of a chronically present pain. In self-reporting, reported crisis events also may be associated flare-up of a chronic pain or neuropathy subjectively perceived as a VOC [35].

Both the development and alleviation of chronic pain would not necessarily be subject to the similar mechanisms as singular VOCs, or even be subject to resolution without an ongoing treatment. To address the uncertainty in the “correct” time-to-resolution, the analysis was performed with events separated into sets with progressively increasing TTR durations (up to 7 days, 14 days, 28 days, and up to 48 days). Shorter TTR durations may better associate with the actual resolution of a VOC, a consideration that is supported by the data presented above. Sample size limitations determined the minimum “feasible” duration of self-reported TTR at ≤ 7 days (TTR_1_).

## Conclusions

This is the first study to expand on findings from the PISCES study [37]. We captured the time to resolution from a VOC using an electronic patient-reported outcomes device and acquired parallel serial biomarker measurements. This study also evaluated both cell functional and blood-based inflammatory biomarkers for longitudinal assessment of clinical baseline. The results from this study suggest that the TTR at ≤ 7 days (TTR_1_), a timeframe consistent with acute VOC, can be predicted, in part, by FA-WB-VCAM and FA-WB-Psel with the predictive capability potentially increased further when used in combination with blood-based inflammatory biomarkers. Multi-parametric modeling identified several combinations of these biomarkers with the potential to highly predict VOC duration. Relative impact of biomarkers predictive of TTR may be indicative of key pathophysiological drivers of VOC and its resolution. Longer perceived TTRs are more likely to be associated with chronic pain or neuropathies. Thus, a correlation of the actual TTR duration with the one predicted by the model, could be used, when validated, as a metric to proactively assess the etiology of the pain event. Biomarker-based differentiation between acute VOC and a chronic pain flare-up could enable better and more precise patient care. These results should be used in further targeted investigations to better understand the etiology of VOC resolution and associated methods for predicting clinical outcomes. Incorporation of validated blood-based biomarkers in clinical care could enable better management of VOC through more patient-centristic interventions, improving quality of life and reducing healthcare costs.

## Data Availability

All data produced in the present study are available upon reasonable request to the authors

## Acknowledgements

The authors gratefully acknowledge all subjects and individuals who contributed to the study. We thank David Beidler and Debra D. Pittman for their role in the design and initiation of the study. We also thank Sanguine Biosciences, Clinical Ink, and CRF Health for their expertise and efforts to make this study possible.

## Bibliography

[1] Kato GJ, Piel FB, Reid CD, Gaston MH, Ohene-Frempong K, Krishnamurti L, Smith WR, Panepinto JA, Weatherall DJ, Costa FF and Vichinsky EP. Sickle cell disease. Nature Reviews Disease Primers 2018; 4: 18010.

[4] Bakshi N, Gillespie S, McClish D, McCracken C, Smith WR and Krishnamurti L. Intraindividual pain variability and phenotypes of pain in sickle cell disease: a secondary analysis from the Pain in Sickle Cell Epidemiology Study. Pain 2021;

[5] Smith WR, Penberthy LT, Bovbjerg VE, McClish DK, Roberts JD, Dahman B, Aisiku IP, Levenson JL and Roseff SD. Daily assessment of pain in adults with sickle cell disease. Ann Intern Med 2008; 148: 94–101.

[6] Shapiro BS, Dinges DF, Orne EC, Bauer N, Reilly LB, Whitehouse WG, Ohene-Frempong K and Orne MT. Home management of sickle cell-related pain in children and adolescents: natural history and impact on school attendance. Pain 1995; 61: 139–144.

[7] Manwani D and Frenette PS. Vaso-occlusion in sickle cell disease: pathophysiology and novel targeted therapies. Blood 2013; 122: 3892–3898.

[8] Mayadas TN, Johnson RC, Rayburn H, Hynes RO and Wagner DD. Leukocyte rolling and extravasation are severely compromised in P selectin-deficient mice. Cell 1993; 74: 541–554.

[9] Lawrence MB and Springer TA. Leukocytes roll on a selectin at physiologic flow rates: Distinction from and prerequisite for adhesion through integrins. Cell 1991; 65: 859–873.

[10] Matsui NM, Borsig L, Rosen SD, Yaghmai M, Varki A and Embury SH. P-selectin mediates the adhesion of sickle erythrocytes to the endothelium. Blood 2001; 98: 1955–1962.

[11] Setty BN and Stuart MJ. Vascular cell adhesion molecule-1 is involved in mediating hypoxia-induced sickle red blood cell adherence to endothelium: potential role in sickle cell disease. Blood 1996; 88: 2311–2320.

[12] White J, Lancelot M, Sarnaik S and Hines P. Increased erythrocyte adhesion to VCAM-1 during pulsatile flow: Application of a microfluidic flow adhesion bioassay. Clin Hemorheol Microcirc 2015; 60: 201–213.

[13] White J, Callaghan MU, Gao X, Liu K, Zaidi A, Tarasev M and Hines PC. Longitudinal assessment of adhesion to vascular cell adhesion molecule-1 at steady state and during vaso-occlusive crises in sickle cell disease. Br J Haematol 2022; 196: 1052–1058.

[14] Pittman DD, Hines PC, Beidler D, Rybin D, Frelinger AL, Michelson AD, Liu K, Gao X, White J, Zaidi AU, Charnigo RJ and Callaghan MU. Evaluation of Longitudinal Pain Study in Sickle Cell Disease (ELIPSIS) by patient-reported outcomes, actigraphy, and biomarkers. Blood 2021; 137: 2010–2020.

[15] Ballas SK. The sickle cell painful crisis in adults: phases and objective signs. Hemoglobin 1995; 19: 323–333.

[16] Arigliani M, Zheng S, Ruiz G, Chakravorty S, Bossley CJ, Rees D and Gupta A. Comparison of pulse oximetry and earlobe blood gas with CO-oximetry in children with sickle cell disease: a retrospective review. BMJ Paediatr Open 2020; 4: e000690.

[17] Nouraie M, Lee JS, Zhang Y, Kanias T, Zhao X, Xiong Z, Oriss TB, Zeng Q, Kato GJ, Gibbs JS, Hildesheim ME, Sachdev V, Barst RJ, Machado RF, Hassell KL, Little JA, Schraufnagel DE, Krishnamurti L, Novelli E, Girgis RE, Morris CR, Rosenzweig EB, Badesch DB, Lanzkron S, Castro OL, Goldsmith JC, Gordeuk VR and Gladwin MT. The relationship between the severity of hemolysis, clinical manifestations and risk of death in 415 patients with sickle cell anemia in the US and Europe. Haematologica 2013; 98: 464–472.

[18] Hines PC, Callaghan MU, Zaidi AU, Gao X, Liu K, White J and Tarasev M. Flow adhesion of whole blood to P-selectin: a prognostic biomarker for vaso-occlusive crisis in sickle cell disease. British Journal of Haematology 2021; 194: 1074–1082.

[19] Keikhaei B, Mohseni AR, Norouzirad R, Alinejadi M, Ghanbari S, Shiravi F and Solgi G. Altered levels of pro-inflammatory cytokines in sickle cell disease patients during vaso-occlusive crises and the steady state condition. Eur Cytokine Netw 2013; 24: 45–52.

[20] Perkins LA, Nyiranshuti L, Little-Ihrig L, Latoche JD, Day KE, Zhu Q, Tavakoli S, Sundd P, Novelli EM and Anderson CJ. Integrin VLA-4 as a PET imaging biomarker of hyper-adhesion in transgenic sickle mice. Blood Adv 2020; 4: 4102–4112.

[21] Dworkis DA, Klings ES, Solovieff N, Li G, Milton JN, Hartley SW, Melista E, Parente J, Sebastiani P, Steinberg MH and Baldwin CT. Severe sickle cell anemia is associated with increased plasma levels of TNF-R1 and VCAM-1. Am J Hematol 2011; 86: 220–223.

[22] Hatzipantelis ES, Pana ZD, Gombakis N, Taparkou A, Tzimouli V, Kleta D, Zafeiriou DJ, Garipidou V, Kanakoudi F and Athanassiou M. Endothelial activation and inflammation biomarkers in children and adolescents with sickle cell disease. Int J Hematol 2013; 98: 158–163.

[23] Odièvre MH, Bony V, Benkerrou M, Lapouméroulie C, Alberti C, Ducrocq R, Jacqz-Aigrain E, Elion J and Cartron JP. Modulation of erythroid adhesion receptor expression by hydroxyurea in children with sickle cell disease. Haematologica 2008; 93: 502–510.

[24] Styles LA, Lubin B, Vichinsky E, Lawrence S, Hua M, Test S and Kuypers F. Decrease of very late activation antigen-4 and CD36 on reticulocytes in sickle cell patients treated with hydroxyurea. Blood 1997; 89: 2554–2559.

[25] Marui N, Offermann MK, Swerlick R, Kunsch C, Rosen CA, Ahmad M, Alexander RW and Medford RM. Vascular cell adhesion molecule-1 (VCAM-1) gene transcription and expression are regulated through an antioxidant-sensitive mechanism in human vascular endothelial cells. J Clin Invest 1993; 92: 1866–1874.

[26] Verger E, Schoëvaërt D, Carrivain P, Victor JM, Lapouméroulie C and Elion J. Prior exposure of endothelial cells to hydroxycarbamide alters the flow dynamics and adhesion of sickle red blood cells. Clin Hemorheol Microcirc 2014; 57: 9–22.

[27] Hattori R, Hamilton KK, Fugate RD, McEver RP and Sims PJ. Stimulated secretion of endothelial von Willebrand factor is accompanied by rapid redistribution to the cell surface of the intracellular granule membrane protein GMP-140. J Biol Chem 1989; 264: 7768–7771.

[28] Akgür FM, Zibari GB, McDonald JC, Granger DN and Brown MF. Kinetics of P-selectin expression in regional vascular beds after resuscitation of hemorrhagic shock: a clue to the mechanism of multiple system organ failure. Shock 2000; 13: 140–144.

[29] Straley KS and Green SA. Rapid transport of internalized P-selectin to late endosomes and the TGN: roles in regulating cell surface expression and recycling to secretory granules. J Cell Biol 2000; 151: 107–116.

[30] Toledo SLO, Guedes JVM, Alpoim PN, Rios DRA and Pinheiro MB. Sickle cell disease: Hemostatic and inflammatory changes, and their interrelation. Clin Chim Acta 2019; 493: 129–137.

[31] Sarray S, Saleh LR, Lisa Saldanha F, Al-Habboubi HH, Mahdi N and Almawi WY. Serum IL-6, IL-10, and TNFα levels in pediatric sickle cell disease patients during vasoocclusive crisis and steady state condition. Cytokine 2015; 72: 43–47.

[32] Uwaezuoke SN, Ayuk AC, Ndu IK, Eneh CI, Mbanefo NR and Ezenwosu OU. Vaso-occlusive crisis in sickle cell disease: current paradigm on pain management. Journal of pain research 2018; 11: 3141–3150.

[33] Brandow AM, Zappia KJ and Stucky CL. Sickle cell disease: a natural model of acute and chronic pain. Pain 2017; 158 Suppl 1: S79–s84.

[34] Robieux IC, Kellner JD, Coppes MJ, Shaw D, Brown E, Good C, O’Brodovich H, Manson D, Olivieri NF, Zipursky A and et al. Analgesia in children with sickle cell crisis: comparison of intermittent opioids vs. continuous intravenous infusion of morphine and placebo-controlled study of oxygen inhalation. Pediatr Hematol Oncol 1992; 9: 317–326.

[35] van Tuijn CF, van Beers EJ, Schnog JJ and Biemond BJ. Pain rate and social circumstances rather than cumulative organ damage determine the quality of life in adults with sickle cell disease. Am J Hematol 2010; 85: 532–535.

[36] Panepinto JA, Brousseau DC, Hillery CA and Scott JP. Variation in hospitalizations and hospital length of stay in children with vaso-occlusive crises in sickle cell disease. Pediatr Blood Cancer 2005; 44: 182–186.

[37] Smith WR, Bovbjerg VE, Penberthy LT, McClish DK, Levenson JL, Roberts JD, Gil K, Roseff SD and Aisiku IP. Understanding pain and improving management of sickle cell disease: the PiSCES study. J Natl Med Assoc 2005; 97: 183–193.

